# Safety and efficacy of inhaled mannitol in patients with cystic fibrosis: Metanalysis

**DOI:** 10.1101/2025.03.02.25323206

**Authors:** Madho Mal, Nayanika Tummala, Laila Shalabi, Mayank Jha, Mian Zahid Jan Kakakhel, Sahira Suman, Rekha Rani, Twinkle Sonia, Parkash Bachani, Love Kumar

**Affiliations:** Liaquat University of Medical and Health Science Jamshoro; GITAM Institute of Medical Science and Research, Hyderabad; Faculty of Medicine, Gharyan University, Gharyan, Libya; Department of Medicine, Government Medical College and New Civil Hospital, Surat, Gujarat, India; Rehman Medical College, Peshawar, Pakistan; Riphah International University, Pakistan; Stamford Hospital, CT, USA; Baptist Hospital of South Texas Beaumont Texas; Jannah Medical and Dental College Karachi Pakistan; Marshall University School of Medicine, All India Institute of Medical Sciences (AIIMS), Jodhpur, India

**Keywords:** Cystic fibrosis, cystic fibrosis diagnosis, mannitol administration and dosage, mannitol adverse effects, Osmitrol, Bronchitol

## Abstract

**Background:** Cystic fibrosis is a multi-system autosomal recessive disorder with considerable mortality and morbidity associated with respiratory manifestations. Due to thick, viscous mucus production, which causes airway obstruction and subsequent respiratory infections, multiple drugs are being used to clear the secretions. Inhaled mannitol is one treatment regime that acts by increasing mucociliary clearance; however, its exact mechanism of action is unknown. Consequently, it has produced effective results in multiple studies.

**Aims:** To determine whether inhaled mannitol is efficacious and safe in treating cystic fibrosis. To assess and compare the therapeutic advantages and the adverse effects.

**Methods:** This systematic review followed the Cochrane Collaboration’s guidelines and the PRISMA format. Relevant literature was derived from databases like Medline, the Cochrane Central Register of RCTs, PubMed Central, and clinicaltrials.gov. Recent research conduction: February 29th, 2022. Selection criteria included all randomized control trials assessing diagnosed CF patients, evaluating the efficacy of inhaled mannitol, and comparing it with a placebo. The authors independently assessed studies for inclusion, carried out data extraction, and assessed the risk of bias in the included studies.

**Results:** A total of 6 RCTs were included, containing --- participants with diagnosed CF. The outcome measure was an increase in FEV1 after inhalation of dry mannitol. Additionally, it was also found to be associated with adverse events like cough (p-value <0.051, OD ratio 1.665), hemoptysis (p-value < 0.469, OD ratio 1.311), and chest discomfort. However, these symptoms were managed conservatively, and ultimately, the intervention proved tolerable.

**Conclusion:** Inhaled mannitol has been shown to improve pulmonary function and mucociliary clearance, relieving some of the symptoms of Cystic fibrosis. However, there is no evidence of improving the overall quality of life and general health. This review suggests that mannitol could be considered as a treatment for cystic fibrosis.

## Introduction

Cystic fibrosis (CF) is a multi-systemic autosomal recessive disease caused by a defect in the expression of CFTR protein, i.e., chloride channel present in the apical membrane of respiratory, digestive, reproductive, and sweat gland epithelium (1). It clinically manifests in a multi-system manner, including a pertussis-like cough, severe dyspnea, recurrent otitis media, sinusitis, polyps, malnutrition, diarrhea, edema, and much more. The main comorbidity in adulthood is respiratory involvement, with the presence of bronchiectasis, chronic bronchial infection, and airflow obstruction (2). Defective CFTR results in thick and sticky mucus obstructing the pathways (3), leading to serious lung infections, especially by pseudomonas (4).

Previously, the treatment focus was on symptomatic improvement and complication prevention, but recently, protein rectifiers are being studied, which are claimed to correct underlying structural and functional abnormalities (4). Pulmonary ventilation is improved with adequate hydration of the patient, mucolytics, bronchodilators, and anti-inflammatory drugs (1).

Among the latest treatment regimens is the use of inhaled mannitol, also known as bronchitis. It is a naturally occurring sugar alcohol, which, when inhaled, creates a change in the osmotic gradient, leading to the movement of water into the CF airway, hydrating the airway surface liquid and enhancing mucociliary clearance (5). Another suggested mechanism of action is stimulating the release of mediators that increase ciliary beat frequency (6, 7). Regardless of the exact mechanism by which it is achieved, mannitol appears to increase mucociliary clearance in people with CF (8, 9, 10).

Regardless, mannitol has shown some positive findings. Two near-identical, randomized, multicenter, double-blind, controlled, parallel-group phase 3 studies investigated the safety and efficacy of inhaled mannitol in subjects with CF (11, 12). They showed significant improvements in FEV_1_ within 6 weeks of treatment commencement (13, 14).

This meta-analysis aims to evaluate the efficacy and safety of inhaled mannitol in patients with CF and provides a comprehensive and compact review of both the advantages and disadvantages of this treatment.

## Method

We followed the recommendations of the PRISMA statement (13).

### Safety and efficacy of inhaled mannitol in patients with cystic fibrosis

There are almost 30,000 patients with cystic fibrosis in the United States. Around half of these patients are aged 18 or older. The children suffering from cystic fibrosis are mostly asymptomatic, so the disease burden is low. The two most common complications associated are pancreatic insufficiency and a decline in lung function.]

1. short description of the medical problem and its importance in terms of disease burden and policy relevance.
2. Describe the intervention under consideration with all possible alternatives. This may also include key features of intervention, such as its clinical presentation and management.
3. The efficacy of the intervention/therapy on the target population
4. Justification to carry out the systematic review. It should identify how much is already known and what needs to be known

This systematic review followed the Cochrane Collaboration’s guidelines and the Preferred Reporting Items for Systematic Reviews and Meta-Analyses (PRISMA) format (15).

### Data Source and Search Strategy

Three authors independently retrieved relevant literature from databases such as Medline, the Cochrane Central Register of RCTs, PubMed Central, and clinicaltrials.gov. The most recent research was conducted on February 29th, 2022.

Our search strategy was extensive and focused on the keywords listed in Table 1. To make a search strategy, we have used Mesh Terms, synonyms, and some common brand names of inhaled mannitol. Additionally, we conducted a manual search of clinical trials to ensure a comprehensive search. Using EndNote software, we retrieved 102 publications, 20 of which were excluded due to duplication and another 28 due to irrelevance. Additionally, 32 of the remaining 54 articles were excluded based on our exclusion criteria. We were unable to retrieve five articles. Finally, we included seven publications in our final review.

**Table:** Summary of Key Randomized Controlled Trials (RCTs) on Mannitol Therapy.

| AUTHORS | STUDY DESIGN | NUMBER OF PARTICIPANTS | INTERVENTION | MEAN AGE | GENDER | BMI (Mean (SD)) | FEV 1 |
| --- | --- | --- | --- | --- | --- | --- | --- |
| Bilton et al | RCT | Mannitol: 177<br>Control: 118 | 400 mg mannitol (n = 177) vs 50 mg mannitol (n = 118) | Mean (SD): 23 (11.3) years<br>Aged < 18 years: n = 105 | 55.3% male (n = 163)<br>44.7% female (n = 132) | Mannitol: 21.07 (3.99) kg/m2;<br>Control: 20.38 (3.59) kg/m2 | Mean (SD): 62.0 (16.3) % predicted |
| Aitken et al | RCT | Mannitol: 184<br>Control: 121 | 400 mg mannitol (n = 184) vs 50 mg mannitol (n = 121) | Mean: 20 years<br>Range: 6 - 53 years<br>Aged < 18 years: n = 154 | 51.5% male (n = 157)<br>48.5% female (n = 148) | Mannitol: 20 (4.1) kg/m2;<br>Control: 19.8 (3.7) kg/m2 | Mean (SD) 63.8 (15.9) % predicted |
| Flume et al | RCT | Mannitol: 209<br>Control: 214 | 400 mg mannitol (n = 209) vs 50 mg mannitol (n = 214) | Mean (SD): 26.8 (7.63) years | 53% male (n = 224)<br>47% female (n = 199) | - | Mean (SD): 63.9 (13.54)% predicted |
| Jaques et al | RCT | Mannitol: 21<br>Control: 18 | 420 mg mannitol (n = 21) vs 30 mg mannitol (n = 18) | Mean: 19.1 years<br>Range: 8 - 48 years | 41% male (n = 16)<br>59% female (n = 23) | Mannitol: 19.9 (4.2) kg/m2;<br>Control: 19.2 (3.3) kg/m2 | Mannitol group Mean (SD) 64.9 (13.0) % predicted<br>Control group: Mean (SD) 64.4 (11.8) % predicted |
| Boeck et al | RCT | Mannitol: 177<br>Control: 118 | 400mg mannitol vs control<br>Baseline | Mean age (SD): | 40% male (n = 37) | - | Mean (SD): 72.23 |
|  |  |  | characteristics reported for 92 participants<br>Results reported for 87 participants | 12.0 (3.0) years<br>Range 6 to 17 years | 60% female (n = 55) |  | (11.6) % predicted |

### Search Strategy

#### Cystic Fibrosis

Cystic Fibrosis Complications, Cystic Fibrosis Cytology, Cystic Fibrosis Diagnosis, Cystic Fibrosis Diagnosis, Cystic Fibrosis Drug Therapy, Cystic Fibrosis Genetics, Cystic immunology, Cystic Fibrosis Pathology, Cystic Fibrosis Physiology, Cystic Fibrosis Rehabilitation, CF, fibrocystic disease of pancreas, mucoviscidosis, pancreatic fibrosis, CFTR

#### Mannitol

Mannitol administration and dosage, Mannitol adverse effects, Mannitol immunology, Mannitol metabolism, Mannitol organization and administration, Mannitol pharmacokinetics, Mannitol pharmacology, Mannitol therapeutic use, Mannitol therapy, Mannitol toxicity, Osmitrol, Bronchitol, d-Mannitol, mannite, manna sugar, an osmotic diuretic, Osmitrol in table 01.

### Inclusion and Exclusion Criteria

Three authors reviewed all relevant publications independently and excluded articles that did not meet the inclusion and exclusion criteria. Following that, the results were compared, and seven articles included were reviewed to ensure they met our inclusion criteria: 1. Randomized controlled trials (RCT). 2. Diagnosed Cystic fibrosis patients. 3. Studies focusing on evaluating the efficacy and safety of inhaled mannitol. 4. Studies focusing on inhaled mannitol and placebo comparison. Studies available in the English language only. 6. Latest published results. The following criteria were used to determine exclusion. 1. Studies published in any other language. 2. Studies including patients with FEV_1_ greater than 90% or less than 30%. 3. Results of open labeled trails. 4. Preliminary results of trials.

### Data Extraction and Quality Assessment

We started extracting data from randomized control trials after selecting studies. Two independent authors were tasked with the responsibility of developing an extraction sheet. The following information was extracted: the RCT number, the first author, the date of publication, the number of participants, the intervention, the inclusion and exclusion criteria, and the primary and secondary outcomes. Both authors’ extraction sheets were compared and finalized.

The risk of bias was assessed using the Cochrane Risk of Bias 2 tool. Five domains were used to evaluate each trial. 1. Process of randomization. 2. Bias resulting from the existence of a period of carryover effects. 3. Incomplete outcome data. 4. Data collection on outcome variables. 5. Reporting of a single result. Each included study was evaluated for quality by two independent authors. Disagreements were resolved using the RoB2 discrepancy tool. Additionally, each trial was approved by the country’s or institution’s ethical review board, and all participants signed an informed consent form.

### Outcome

The efficacy and safety of inhaled mannitol in the treatment of cystic fibrosis were evaluated. To assess efficacy, changes in FEV1 and FVC are compared between the mannitol and placebo groups. The majority of studies used 400mg of inhaled mannitol as an intervention and 50mg of mannitol as a placebo. The safety of mannitol was determined by examining the adverse effects and the number of patients who withdrew from trials due to adverse effects.

## Result

The analysis is based on five studies (Table 02) that evaluated the effect of mannitol drug’s adverse effect cough in cystic fibrosis patients. In each study, patients were randomly assigned to either mannitol or placebo, and the researchers assessed the patients’ cough after treatment.

**Table 02:**
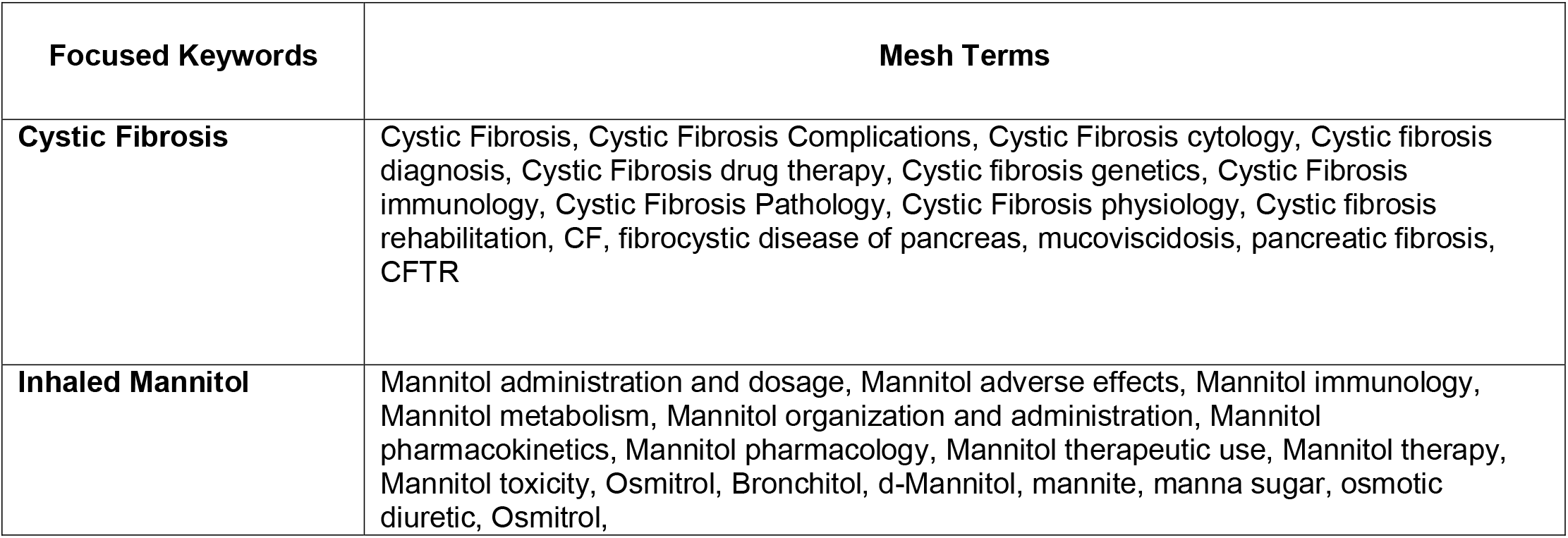
Search strategy and MESH Terms.

The odds ratio is 1.665. Corresponding p-value of < 0.051. We reject the null hypothesis, and this means the drug does decrease cough in the universal populations, which are comparable to those in the analysis. Summarized in Figure (2)

**Figure 01.**
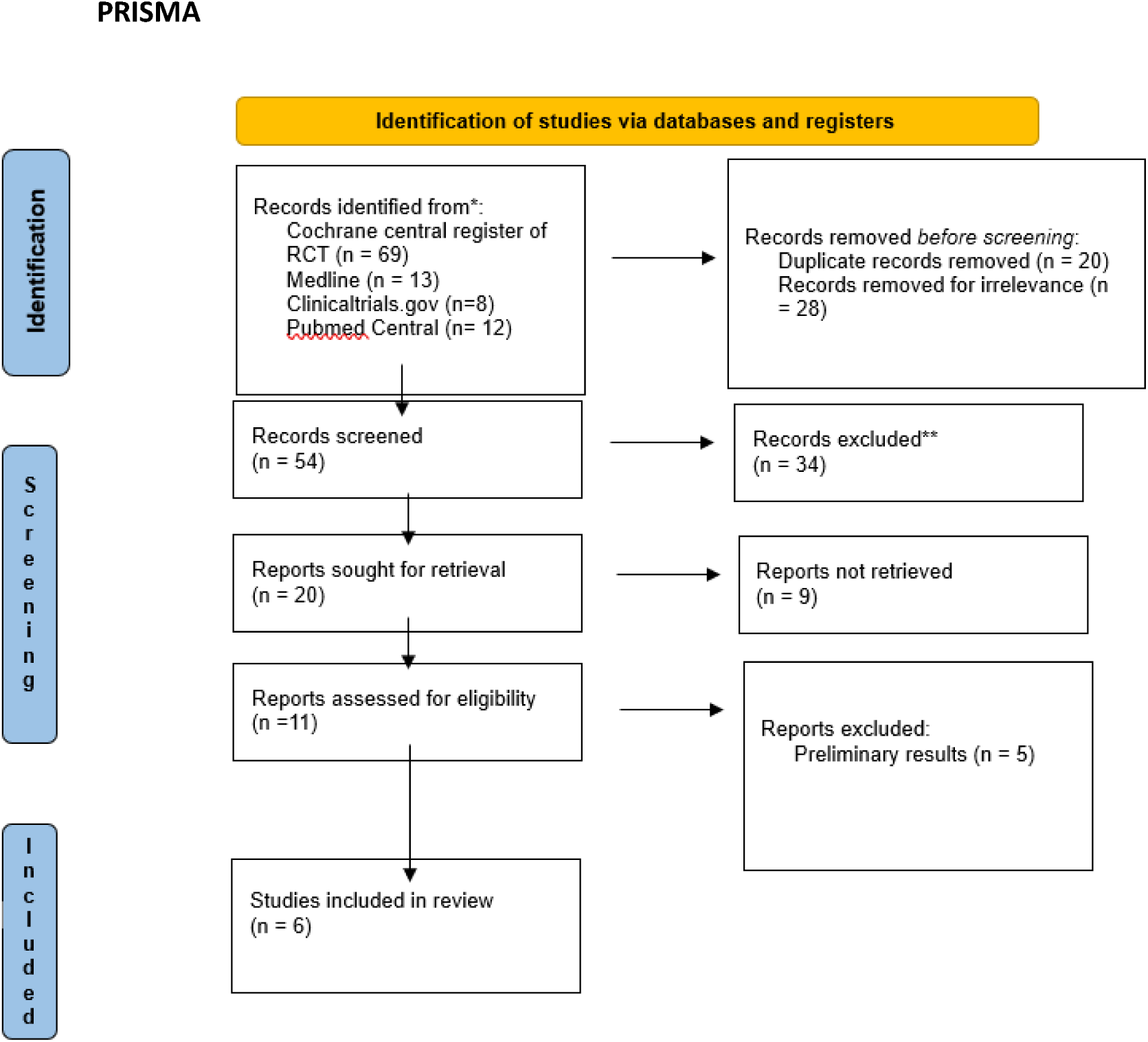

**Figure 02.**
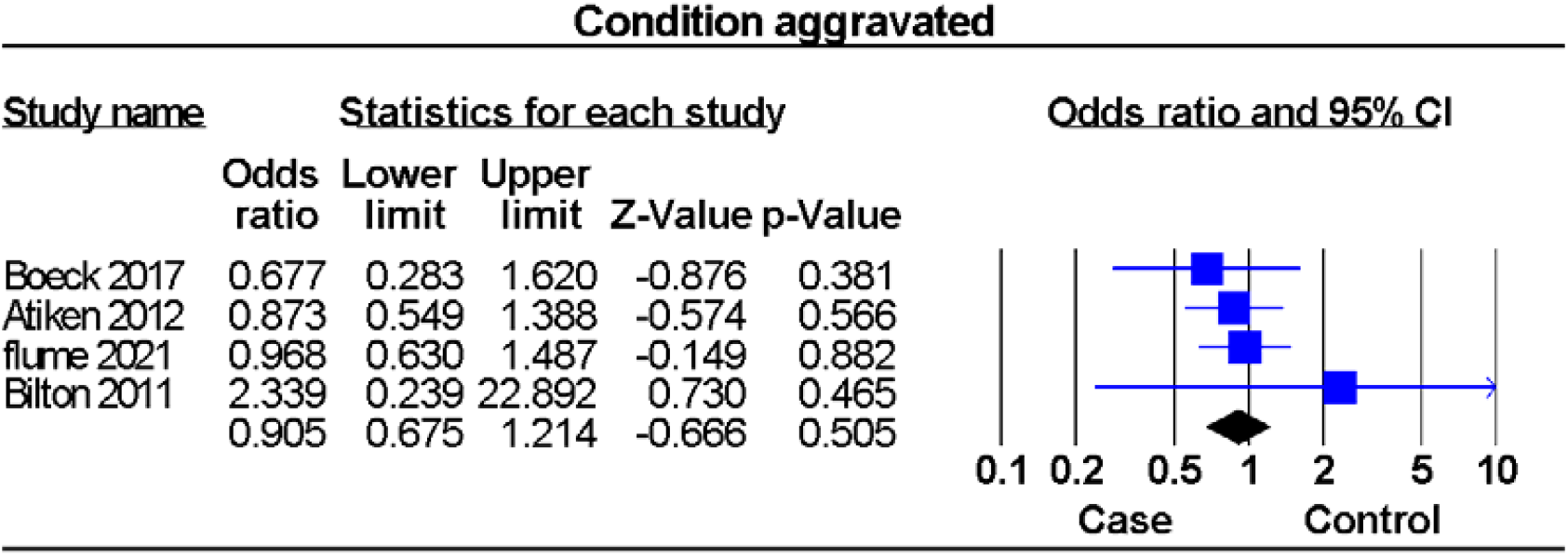

The Q-value is 6.634 with 4 degrees of freedom and p=0.157. We accept the null hypothesis that the true effect size is identical in all the studies. The I2 statistic is 39%, the variance of true effects (T2) is 0.128, and the standard deviation of true effects (T) is 0.357.

The sensitivity of one study removed no effect was shown for a specific study, figure In supplement (1)

In publication by Egger’s Test In this case the intercept (B0) is 0.91909, 95% confidence interval (−4.53751, 6.37568), with t=0.53604, df=3. The 1-tailed p-value is 0.31456, and the 2-tailed p-value is 0 62913.Figure In supplement (1)

The analysis is based on four studies that evaluated the effect of mannitol drug’s adverse effect on headaches in cystic fibrosis patients. In each study, patients were randomly assigned.

The odds ratio is 0.815. Corresponding p-value of < 0.255. We accept the null hypothesis, and this means the drug does not affect headaches in the universal population, which is comparable to those in the analysis. Summarized in Figure (3)

**Figure 03.**
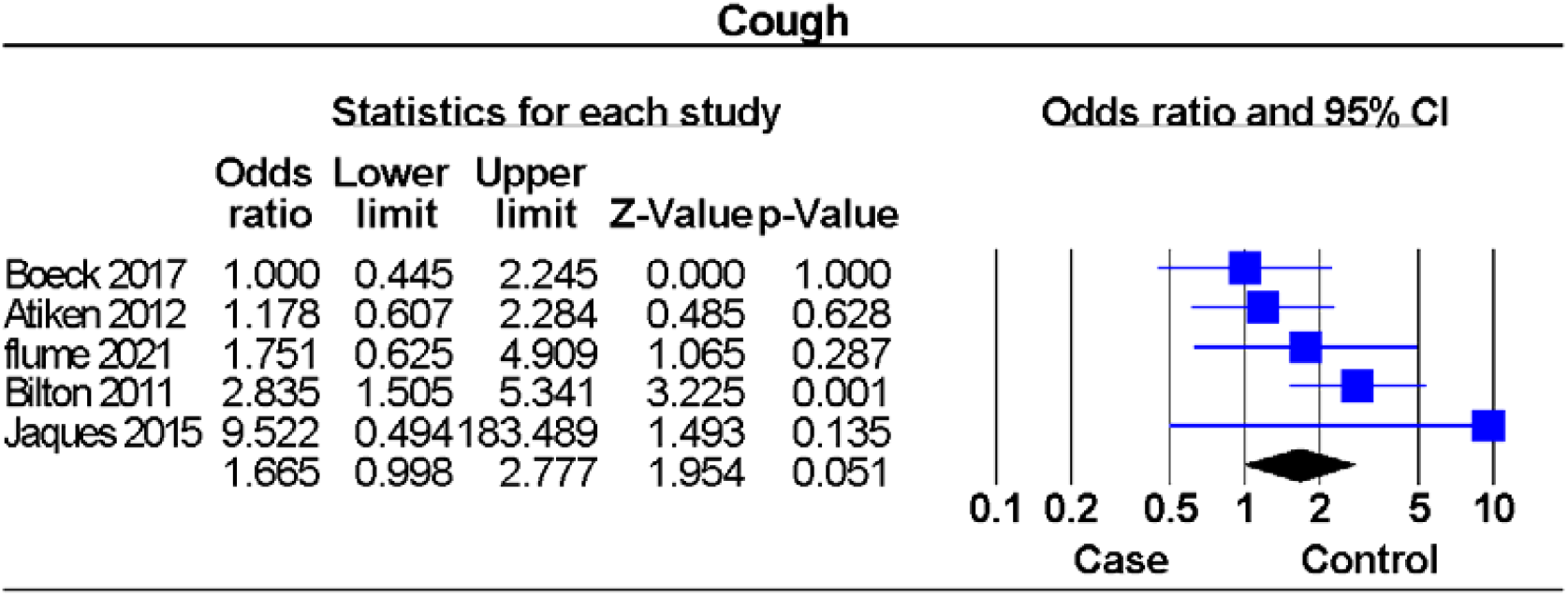

The Q-value is 23.636 with 6 degrees of freedom and p=0.255. We accept the null hypothesis that the true effect size is identical in all the studies.

In publication, and by Egger’s Test of the Intercept In this case the intercept (B0) is 0.44038, 95% confidence interval (−10.54181, 11.42256), with t=0.17253, df=2.

The 1-tailed p-value is 0.43945, and the 2-tailed p-value is 0.87890. Figure in supplement (1)

The analysis is based on four studies that evaluated the effect of mannitol drug and the adverse effect of hemoptysis in cystic fibrosis patients. In each study, patients were randomly assigned.

The odds ratio is 1.311. Corresponding p-value of < 0.469. We accept the null hypothesis, and this means the drug has no effect on hemoptysis in the universal populations, which are comparable to those in the analysis. Summarized in in figure (4)

**Figure 04.**
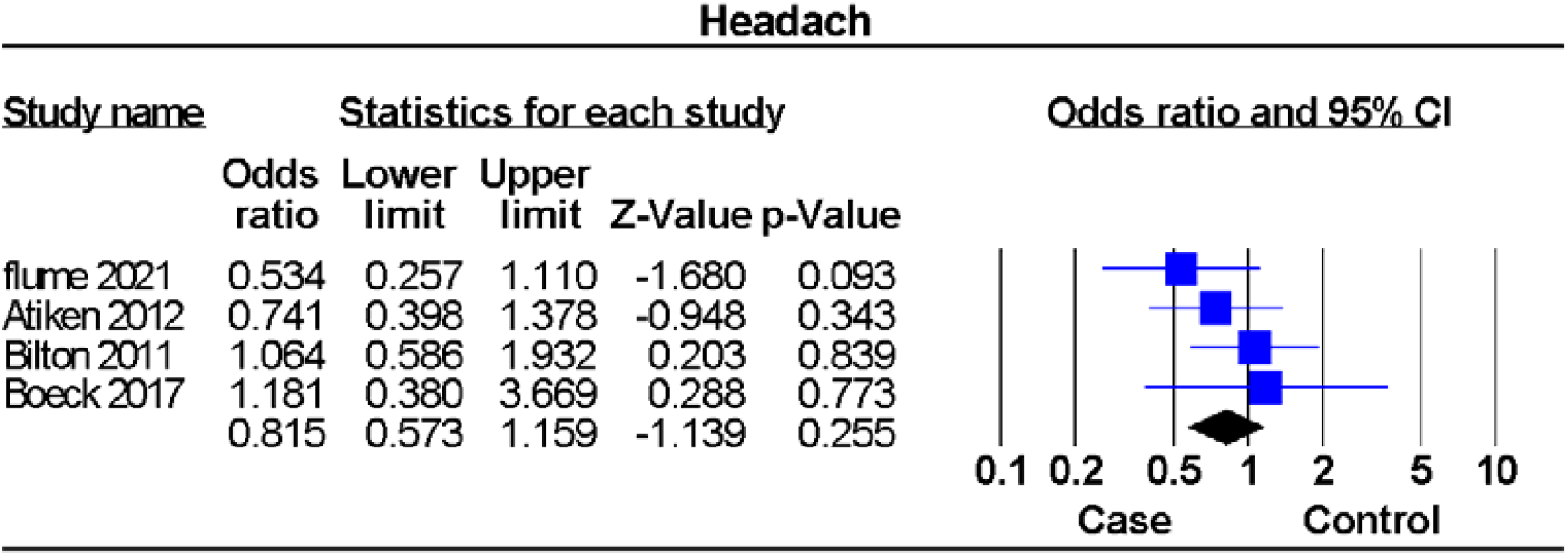

The Q-value is 4.938 with 3 degrees of freedom and p=0.176. We reject the null hypothesis that the true effect size is identical in all the studies. The I2 statistic is 39%, the variance of true effects (T2) is 0.215, and the standard deviation of true effects (T) is 0.464.

The sensitivity of one study removed no effect was shown for a specific study, figure in supplement (1)

In a publication by Egger’s Test of the in this case the intercept (B0) is 0.09937, 95% confidence interval (−11.24757, 11.44631), with t=0.03768, df=2. The 1-tailed p-value is 0.48668, and the 2-tailed p-value is 0.97336. Figure In supplement (1)

The analysis is based on four studies that evaluated the effect of mannitol drug’s adverse effect on nasopharyngitis in cystic fibrosis patients. In each study, patients were randomly assigned to either mannitol or placebo, and the researchers assessed the patients’ nasopharyngitis after treatment.

The odds ratio is 1.171. Corresponding p-value of < 0.483. We accept the null hypothesis, and this means the drug has no effect on nasopharyngitis in the universal populations, which are comparable to those in the analysis. Summarized in in figure (5)

**Figure 05.**
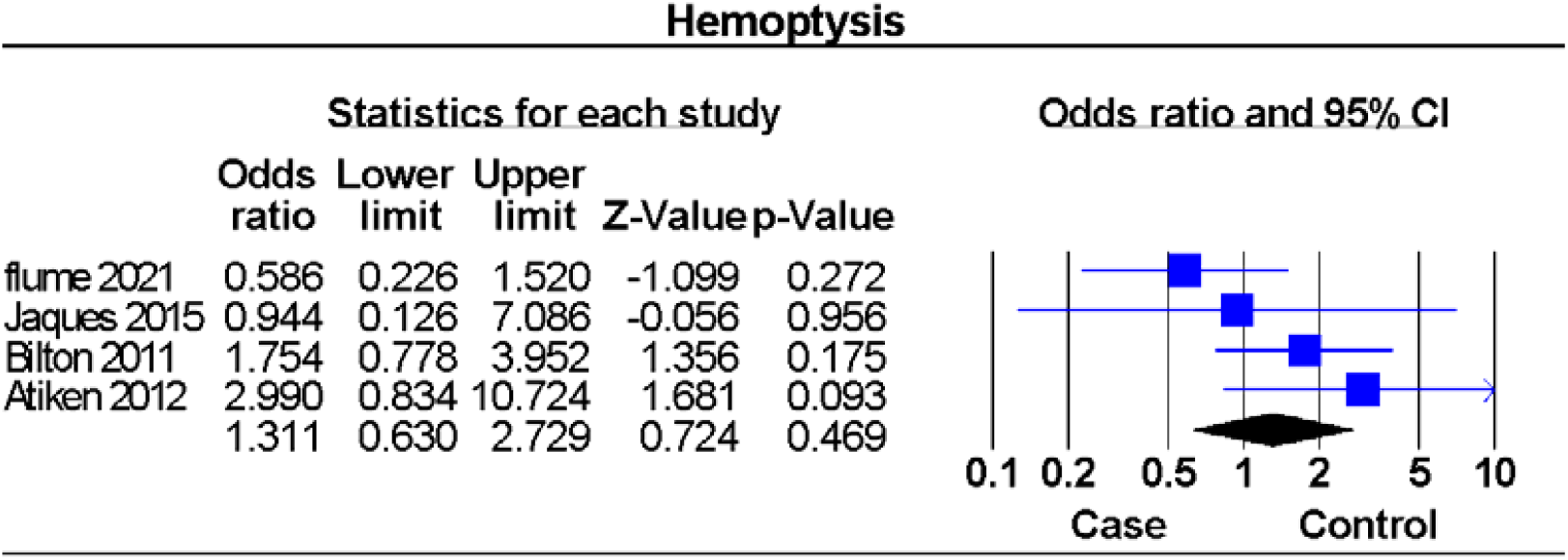

The Q-value is 0.097 with 3 degrees of freedom and p=0.992. We accept the null hypothesis that the true effect size is identical in all the studies.

In publication, and by Egger’s Test of the Intercept In this case the intercept (B0) is -0.35676, 95% confidence interval (−2.51094, 1.79741), with t=0.71258, df=2. The 1-tailed p-value is 0.27501, and the 2-tailed p-value is 0.55002. Figure in supplement (1)

The analysis is based on four studies that evaluated the effect of mannitol drug on adverse effects of Conditions aggravated in cystic fibrosis patients. In each study, patients were randomly assigned.

The odds ratio is 0.905. Corresponding p-value of < 0.505. We accept the null hypothesis, and this means the drug does not affect Conditions aggravated in the universal populations, which are comparable to those in the analysis. Summarized in in figure (7)

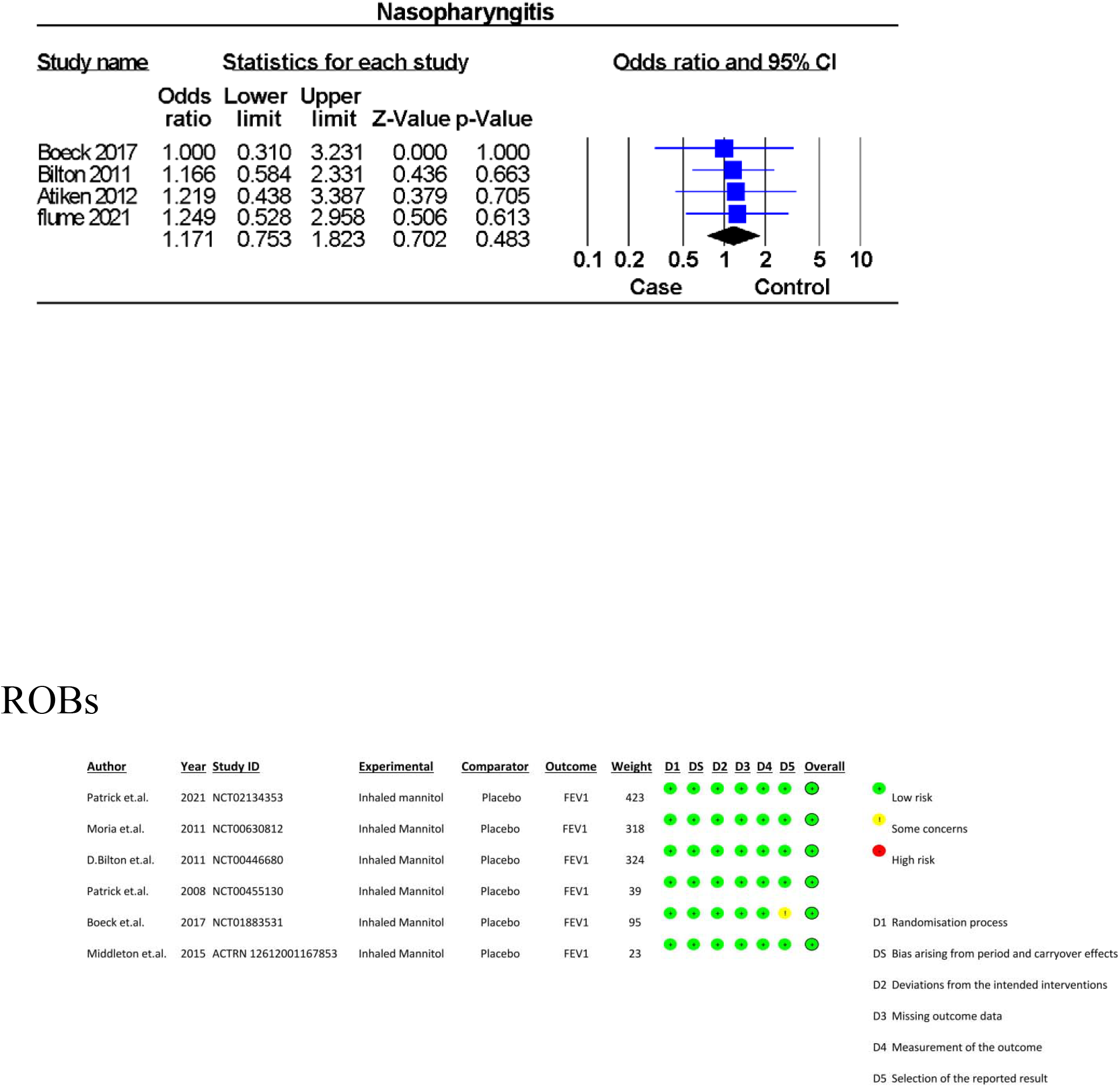

The Q-value is 1.208 with 3 degrees of freedom and p=0.751. We reject the null hypothesis that the true effect size is identical in all the studies.

In publication, and by Egger’s Test of the Intercept In this case the intercept (B0) is 0.39112, 95% confidence interval (−3.10729, 3.88953), with t=0.48103, df=2. The 1-tailed p-value is 0.33899, and the 2-tailed p-value is 0.67798. Figure in supplement (1)

## Discussion

In the meta-analysis, the efficacy and safety of the use of inhalational mannitol is assessed through systematic search, screening, and data extraction with statistical support. Mannitol via the inhalation route has been investigated through various studies in the past as a potential option for treatment for cystic fibrosis patients [1- 3]. The findings in this study revealed significant improvement in pulmonary function evident through a rise in FEV1 among the test groups provided with mannitol compared to control groups [4-8]. These results are in line with previous studies [1,2].

Besides the evidence of benefits of inhalational mannitol, it was found to be associated with adverse effects of cough, hemoptysis, and chest discomfort among the subjects. This was attributed to the route of administration and was able to be managed conservatively and did not result in discontinuation. This supports the existing evidence about the tolerability of the drug for treatment usage. Tolerability showed further improvement in adherence to usage consistently as evident from the trials [4-6]. This forms the main supporting part of treatment success as compliance and tolerability favor better treatment outcomes. The study’s duration was limited to 26-30 weeks, which allowed the investigation of short-term outcomes and side effects. However, to evaluate outcomes and side effects in long-term usage, long-duration studies are needed.

Also, the usage of inhalational mannitol as an adjunct to conventional therapies of Rho DNase, etc., showed sustained benefits in the studies. This supports the fact that it can be a good option for adjunctive therapy along with conventional treatment options. Despite the benefits studied, limitations were faced in these studies as well. Firstly, the modest number of participants involved might have been a factor for positive findings and can be overcome by involving a large scale of subjects from diverse backgrounds to check for outcomes holistically. Secondly, the focus on clinical improvement of pulmonary function tests was shown without analysis of quality of life and exacerbation rates, which would provide further information for improvement of drugs. Incorporating quality-of-life measures in future studies is warranted to get a holistic approach to drug outcomes. Furthermore, the studies included lack of blinding, which can cause bias of information, thus highlighting the need for randomized and double or triple-blinded study design in future trials. In conclusion, the included studies show the efficacy of inhalational powdered mannitol, and improved tolerance is beneficial in drug adherence. Nevertheless, the limitations must be emphasized while framing future research trials and overcome for more supporting evidence.

### Clinical Trial Number

Not applicable

## Data Availability

All data produced are available online at

## Notes

### Competing Interest Statement

The authors have declared no competing interest.

### Funding Statement

This study did not receive any funding

